# The time is ripe to harmonize global acute febrile illness etiologic investigations

**DOI:** 10.1101/2022.05.19.22275321

**Authors:** José Moreira, B. Leticia Fernando-Carballo, Camille Escadafal, Sabine Dittrich, Patrícia Brasil, André M Siqueira

## Abstract

We investigated the impact of considering different case definitions of fever as inclusion criteria among participants enrolling in an acute febrile illness investigation and observed that adopting a subjective assessment of fever regardless of specific fever cut-off value accounted for the diversity of clinical fever phenotypes and did not mischaracterize the febrile population.

## Introduction

Acute febrile illness (AFI) is a common reason for people seeking care and represents a range of infectious and non-infectious diseases, with wide variation by region of the globe [1]. AFI investigations aim to provide national and subregional epidemiology and infectious diseases surveillance and to inform treatment guidelines about empirical and pathogen-specific management algorithms to prioritize interventions and funding. Several AFI investigations have been conducted primarily in Southeast Asia, with few global initiatives in resource-constrained countries [2]. However, there is no standardized approach to conducting such studies, limiting the interpretation and comparability of study findings.

We were involved in a multinational project around fever host-biomarkers [3] and aimed to conduct a pilot study to inform the best criteria for our study site selection. We hypothesized that adopting a subjective or self-reported fever assessment as part of our inclusion criteria would increase the sensitivity threshold for a broad array of febrile clinical phenotypes rather than specifying a cut-off value for fever at presentation. Another hypothesis was that the recent antipyretic intake – largely available over-the-counter-would lower body temperature at arrival at the Emergency Departments (EDs).

## Methods

We did a cross-sectional, study of patients presenting to two urban EDs (Unidade de Pronto Atendimento Manguinhos & Unidade de Pronto Atendimento Rocha Miranda) in Rio de Janeiro, Brazil, between October 28, 2018, and March 29, 2019. The study was conducted following the Declaration of Helsinki and the Brazilian National Ethics Research Committee and was approved in August 2018 (IRB approval no:70984617.9.0000.5262). We screened all consecutive non-severe patients who presented to the EDs over the study period and included those who reported fever ≤ 7 days regardless of temperature measured at the EDs.

Research assistants prospectively collected detailed information and interviewed participants or the primary caregivers to obtain data about the demographics, main chief complaint, vital signs, history of fever, peak measured temperature before arrival at the EDs (temperature maximum), the measured axillary temperature at the EDs, home administration of antipyretics, type/dose/timing of last administration, and tests performed at the EDs. Discharge diagnosis and treatment received after discharge was retrospectively collected by reviewing electronic medical records. The attending physician proposed the discharge diagnosis (not involved in the study), and its appropriateness was not re-evaluated. We used the following definitions: fever was subjectively identified by the participant at arrival at the EDs; antipyretics are mostly over-the-counter drugs that act on reducing body temperature usually with added analgesic effect, and were self-administered by the participants before seeking emergency care; adults those aged ≥ 18 years at recruitment, and non-severe AFI was an illness that was codified as blue or green according to the Manchester Triage System at EDs, which does not require hospital admission. We measured the axillary temperature at triage and estimated the magnitude of defervescence, considering the highest measured self-reported temperature before EDs arrival (Tmax). Suspected infection was defined as the initiation of any antimicrobial after an EDs visit.

## Statistical analysis

Descriptive analysis was performed to characterize the distributions of several variables. A Chi-square test was used to compare categorical variables between study groups. Continuous variables were compared between the study groups using analysis of variance or the Kruskal-Wallis test if those were found to be normally or non-normally distributed, respectively. We dealt with missing data using the listwise deletion method (or complete case analysis).

A logistic regression model was performed to ascertain the effects of recent antipyretic uptake on the likelihood that participants have an overt fever at EDs admission (i.e., ≥ 37.5 °C). A 2-sided *p-value* less than 0.05 was considered statistically significant. All statistical analyses were performed using SPSS software (version 26, IBM, Chicago, Illinois, USA) and Tableau Desktop (version 2020.4.2, Tableau, Seattle, Washington, USA). The primary outcome was to investigate the impact of considering different case definitions of fever as inclusion criteria among febrile patients attending EDs.

## Results

During the study period, we triaged 1551 subjects, and 374 [24.1% (95% CI 22-26)] had a history of fever at arrival at the ED, corresponding to a cumulative incidence of fever at EDs of 24 per 100 triaged patients over the five months. These 374 febrile patients constitute our study population, of whom 248 (66.3%), 115 (30.7%), and 11 (2.9%) had a temperature < 37.5 °C, ≥ 37.5 °C, and no temperature measurement registered at arrival, respectively. The mean age was 30.6 [range: 0-84] years, adults (82.5%) and females (54.8%) predominated (Table). The median peak measured temperature before arrival at the ED was 38.8°C (38.0-39.0), and the median duration of fever was 1 [1-3] days.

**Table.** Baseline characteristics according to the height of temperature mensurated at arrival in febrile patients attending emergency departments in Rio de Janeiro, Brazil, October 2018-March 2019

|  | **Total febrile population** | **History of fever but temperature < 37.5 °C** | **History of fever and temperature ≥ 37.5 °C** | **p-value** |
| --- | --- | --- | --- | --- |
|  | **N= 374** | **N= 248** | **N= 115** |  |
| **Characteristics** |  |  |  |  |
| **Demographics** |  |  |  |  |
| **Age (years), mean (±SD)** | 31 (16) | 32 (15) | 28 (16) | **0.02** |
| **Missing** | 3 (0.8) |  |  |  |
| **Male, n- (%)** | 169 (45.2) | 103 (41.5) | 62 (53.9) | **0.02** |
| **Temp at arrival, (°C), mean (±SD)** | 37.08 (0.9) | 36.55 (0.5) | 38.24 (0.6) | **<0.01** |
| **Missing** | 12 (3.2) |  |  |  |
| **Tmax (°C), mean (±SD)** | 38.7 (0.7) | 38.7 (0.69) | 38.95(0.9) | 0.12 |
| **Missing** | 268 (71.7) |  |  |  |
| **Days of fever, mean (±SD)** | 1.58 (17.4) | 1.45 (21.2) | 1.80 (3.2) | 0.86 |
| **Missing** | 81 (21.7) |  |  |  |
| **Degree of defervescence from Tmax, mean (±SD)** | 1.70 (1.13) | 2.16 (0.86) | 0.78 (1.04) | **<0.01** |
| **Diagnosis and management** |  |  |  |  |
| **Diagnostic testing – n (%)** | 76 (30.2) | 42 (26.1) | 32 (38.1) | 0.05 |
| **Recent antibiotic use – n (%)** | 21 (12.2) | 11 (9.6) | 10 (18.9) | 0.09 |
| **Missing** | 202 (54) |  |  |  |
| **Recent antipyretic use – n (%)** | 249 (66.6) | 169 (87.1) | 76 (90.5) | 0.42 |
| **Missing** | 90 (24.1) |  |  |  |
| **Antibiotic prescription – n (%)** | 131 (53.0) | 75 (47.8) | 54 (65.1) | **0.01** |

SD means standard deviation; Tmax higher temperature self-recorded at home before emergency department visit; Recent antibiotic use was defined as any antibiotic for systemic use started prior to emergency department visit. We excluded antibiotics for systemic use that could also be used for the treatment of tuberculosis in patients with active disease; Recent antipyretic was the use of any over-the counter antipyretic prior to emergency department visit.

Hundred ninety-eight febrile patients had a diagnosis of infection, with upper respiratory tract infection (42.4%) followed by undifferentiated febrile illness (15.7%) and urinary tract infection (14.6%) were the primary febrile syndromes (Supplementary Figure 1). Hundred thirty-one patients were prescribed any antibiotic after EDs visit, which beta-lactams (73/131, 55.7%), quinolones (23/131, 17.5%) and macrolides (16/131,12.2%) were the classes most commonly prescribed. Patients who had temperature ≥ 37.5 °C at presentation were more likely to be diagnosed with an infection [76 (66%) *vs*. 116 (47%), crude OR: 2.2 (95% 1.4-3.5) and were prescribed antibiotic most frequently compared to those with temperature < 37.5 °C [54 (65%) *vs*. 75 (48%), crude OR: 2 (95% 1.1-3.5)].

The site of infection varied by the height of temperature mensurated at arrival at EDs (*p*<0.001). Patients who had a temperature < 37.5 °C at presentation were more likely to be diagnosed with upper respiratory (31.6%) and urinary infections (15.4%) (Supplementary Figure 3). The most frequently etiology diagnostics recorded in those with temperature < 37.5 °C at presentation were acute pharyngitis (20/248, 8%), followed by community-acquired gastroenteritis (19/248, 7.6%), uncomplicated urinary tract infection (16/248, 6.4%), cutaneous abscess (14/248, 5.6%), and community-acquired pneumonia (9/248, 3.6%).

In total, 249/374 (66.6% (95% 61.5-71.3)]) subjects reported self-administering an antipyretic at home within a median of 2h [IQR 2-6] before EDs admission (24.1% of the febrile subjects did not recall prior antipyretics intake). Dipyrone (72.2%) was the antipyretic drug most frequently administered, followed by acetaminophen (14.3%) and ibuprofen (6.3%). Most antipyretic takers consumed one class (85.9%), and the median number of doses taken was 2 [IQR 1-3]. Of these 249 subjects, 167/249 (67%) reported the last dose administered at a mean of 2 [2-5] hours before hospital presentation. Subjects who were treated with an antipyretic drug before arrival did not differ from untreated ones regarding the temperature at arrival at the ED visit (37 ± 0.99 *vs*. 36.8 ± 0.86, *p* = 0.20) or the documented Tmax before the ED arrival (38.8 ± 0.79 *vs*. 38.4 ± 0.60, *p* = 0.29).

The logistic regression model correctly classified 69.1% of the cases (Supplementary Table 1). Males were 2.2 times more likely to exhibit overt fever at presentation than females [aOR: 2.25, 95% CI: 1.19-4.25]. Recent antipyretic intake did not influence the pattern of fever at presentation [aOR:0.81, 95% CI:0.34-1.97].

## Discussion

Our study suggests that considering a subjective/self-reported fever as an inclusion criterion rather than a specific fever cut-off value (i.e., ≥ 37.5 °C) at presentation for AFI etiologic investigation results in a more inclusive study population and account for the diversity of clinical fever phenotypes that are usually seen in clinical practice. Specifically, sixty-six percent of our cohort would not have been included if we restricted our inclusion criteria to a specific febrile cut-off, potentially impacting study recruitment and patient characterization. Almost half (47.8%) of febrile participants with temperature < 37.5 °C at presentation were prescribed at least one antibiotic, and their infections were mainly upper respiratory, justifying their inclusion in AFI studies. Next, our findings suggest that the ubiquitous administration of antipyretics (66.6%) observed in our setting did not influence the febrile presentation of patients attending the EDs evaluated.

We concur with others [2, 4] and argue that there is an urgent need to standardize AFI study protocols to make sure that we can compare results from different studies and pool their results to develop national, regional, or global burden of fever estimates. Such consensus in AFI research would allow better comparisons, translate evidence to practice, and inform treatment policy. A harmonized definition of fever would enable a more robust and consistent capture of different clinical phenotypes, including those conditions where overt fever occurs inconsistently, such as upper respiratory infections and uncomplicated urinary infections. Others [2] have suggested an adapted STRengthening the reporting of OBservational studies in Epidemiology (STROBE) checklist for conducting and reporting AFI research.

In conclusion, we suggest harmonizing the case definition of fever in AFI research for a history of fever regardless of temperature cut-off would enable a complete picture of the different clinical fever phenotypes, thus improving our understanding of the global burden of febrile illness.

## Data Availability

All data produced in the present study are available upon reasonable request to the authors

## Author contributions

JM: writing original draft, visualization, conceptualization. BC: Review and editing. CE: Review and editing. SD: review, editing. PB: conceptualization, writing, editing, supervision. AS: conceptualization, writing, editing, supervision

## Transparency declaration

The authors have no conflict of interest to declare.

## Supplementary Tables

**Supplementary Table 1.**
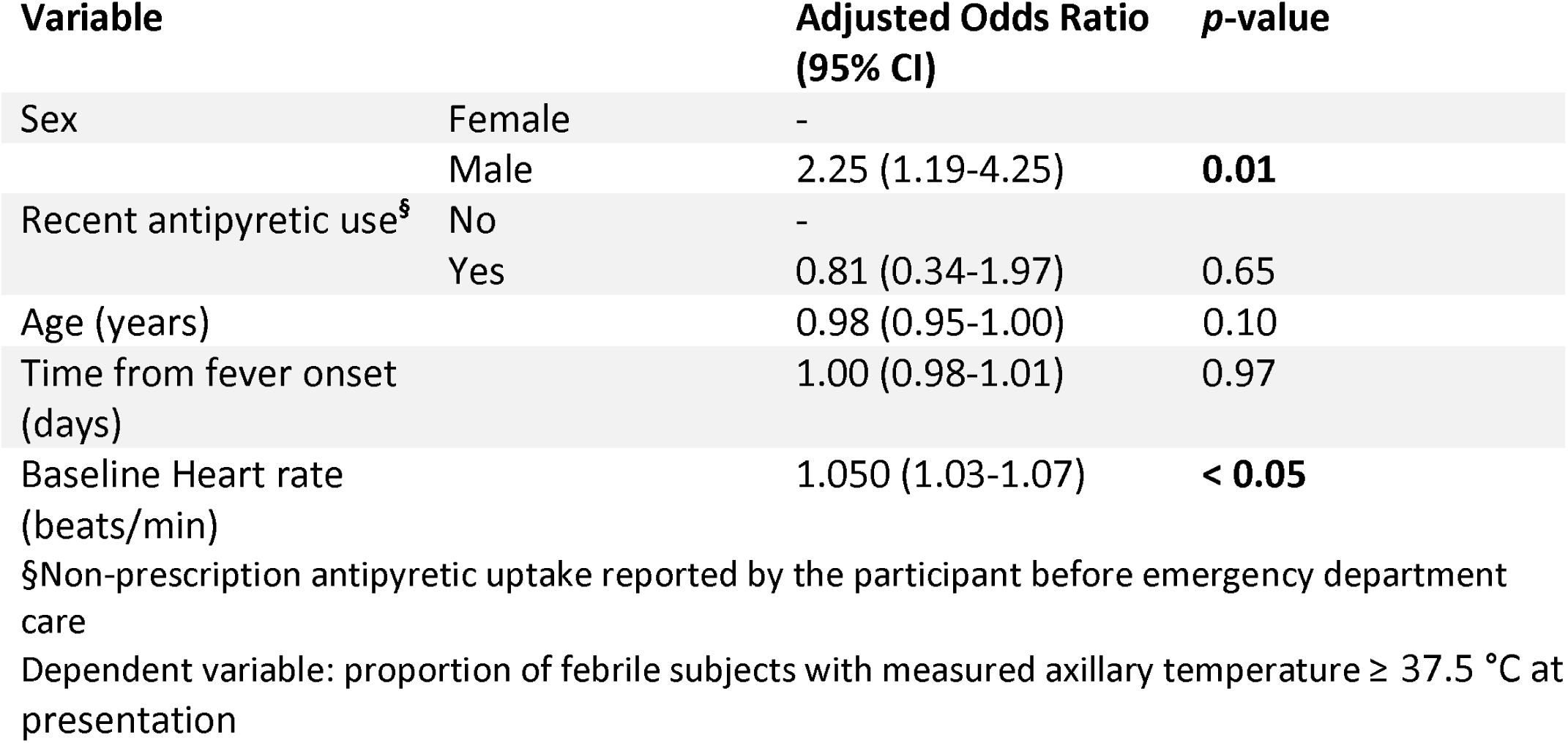
Prediction model for the likelihood of overt fever at presentation in 374 febrile patients attending emergency departments in Rio de Janeiro, Brazil, October 2018 – March 2019.

**Supplementary Table 2.** Diagnosis, febrile syndrome, and antibiotic prescription in febrile patients attending Emergency Departments in Rio de Janeiro, Brazil, October 2018-March 2019

| Presumed diagnosis by febrile syndrome | Number of presentations<br>n/N (%) | Number of patients receiving an antibiotic prescription<br>n/N (%) |
| --- | --- | --- |
| **Gastrointestinal** |  |  |
| Gastroenteritis | 16/17 (94.11) | 7/16 (43.75) |
| Gastritis | 1/17 (5.88) | 1/1 (100) |
| **Upper respiratory tract infection** |  |  |
| Acute tonsillitis | 34/84 (40.47) | 34/34 (100) |
| Acute pharyngitis | 22/84 (26.19) | 5/22 (22.72) |
| Acute sinusitis | 8/84 (9.52) | 6/8 (75) |
| Common cold | 5/84 (5.95) | 0/5 (0) |
| Other upper respiratory tract infections | 15/84 (17.85) | 9/15 (60) |
| **Lower respiratory tract infection** |  |  |
| Acute bronchitis and COPD exacerbations | 3/17 (17.64) | 2/3 (66.66) |
| Community-acquired pneumonia | 9/17 (52.94) | 8/9 (88.88) |
| Influenza | 2/17 (11.76) | 0/2 (0) |
| Other lower respiratory tract infections | 3/17 (17.64) | 3/3 (100) |
| **Skin and soft tissue infection** |  |  |
| Cellulitis and abscesses | 14/19 (73.68) | 14/14 (100) |
| Other infection of the skin and soft tissue | 4/19 (21.05) | 4/4 (100) |
| Bacterial skin infection | 1/19 (5.26) | 1/1 (100) |
| **Urinary tract infection** |  |  |
| Uncomplicated cystitis & pyelonephritis | 28/29 (96.55) | 25/28 (89.28) |
| Other urinary infection | 1/29 (3.44) | 1/1 (100) |
| **Acute undifferentiated febrile illness** |  |  |
| Arboviruses | 5/30 (16.66) | 0/5 (0) |
| Unspecified febrile illness | 6/30 (20) | 4/6 (66.66) |
| Leptospirosis | 2/30 (6.66) | 2/2 (100) |
| Unspecified viral infection | 15/30 (50) | 1/15 (6.66) |
| Other | 1/30 (3.33) | 1/1 (100) |

COPD stands for chronic obstructive pulmonary disease

## Supplementary Figures

**Supplementary Figure 1.**
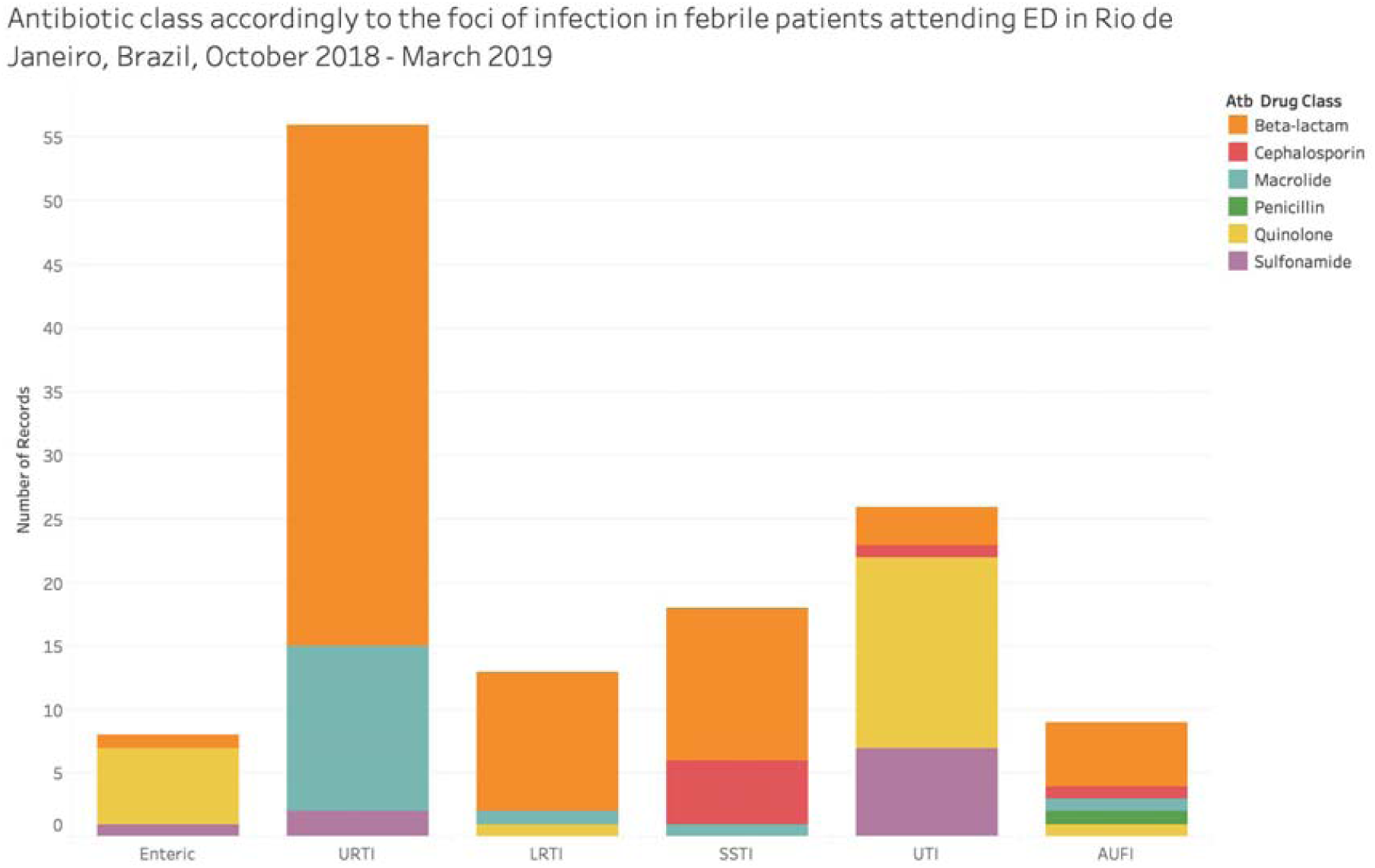
Antibiotic class according to the foci of infection in febrile patients attending Emergency departments in Rio de Janeiro, Brazil, October 2018-March 2019 AUFI stands for acute undifferentiated febrile illness; LRTI lower respiratory tract infection; SSTI skin and soft tissue infection; URTI upper respiratory tract infection; and UTI urinary tract infection

**Supplementary Figure 2.**
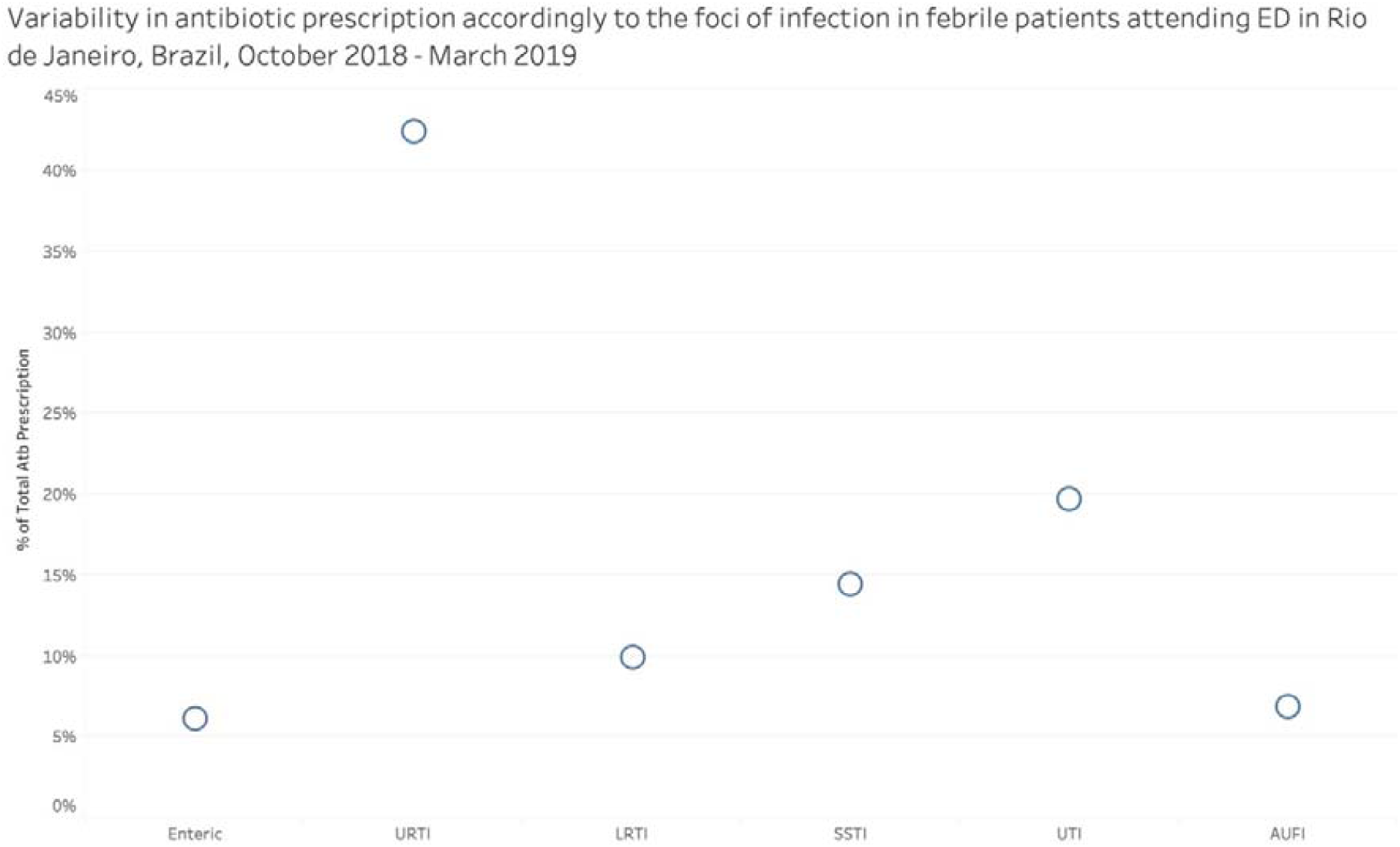
Variability in antibiotic prescription according to the foci of infection in febrile patients attending Emergency departments in Rio de Janeiro, Brazil, October 2018-March 2019 AUFI stands for acute undifferentiated febrile illness; LRTI lower respiratory tract infection; SSTI skin and soft tissue infection; URTI upper respiratory tract infection; and UTI urinary tract infection

**Supplementary Figure 3.**
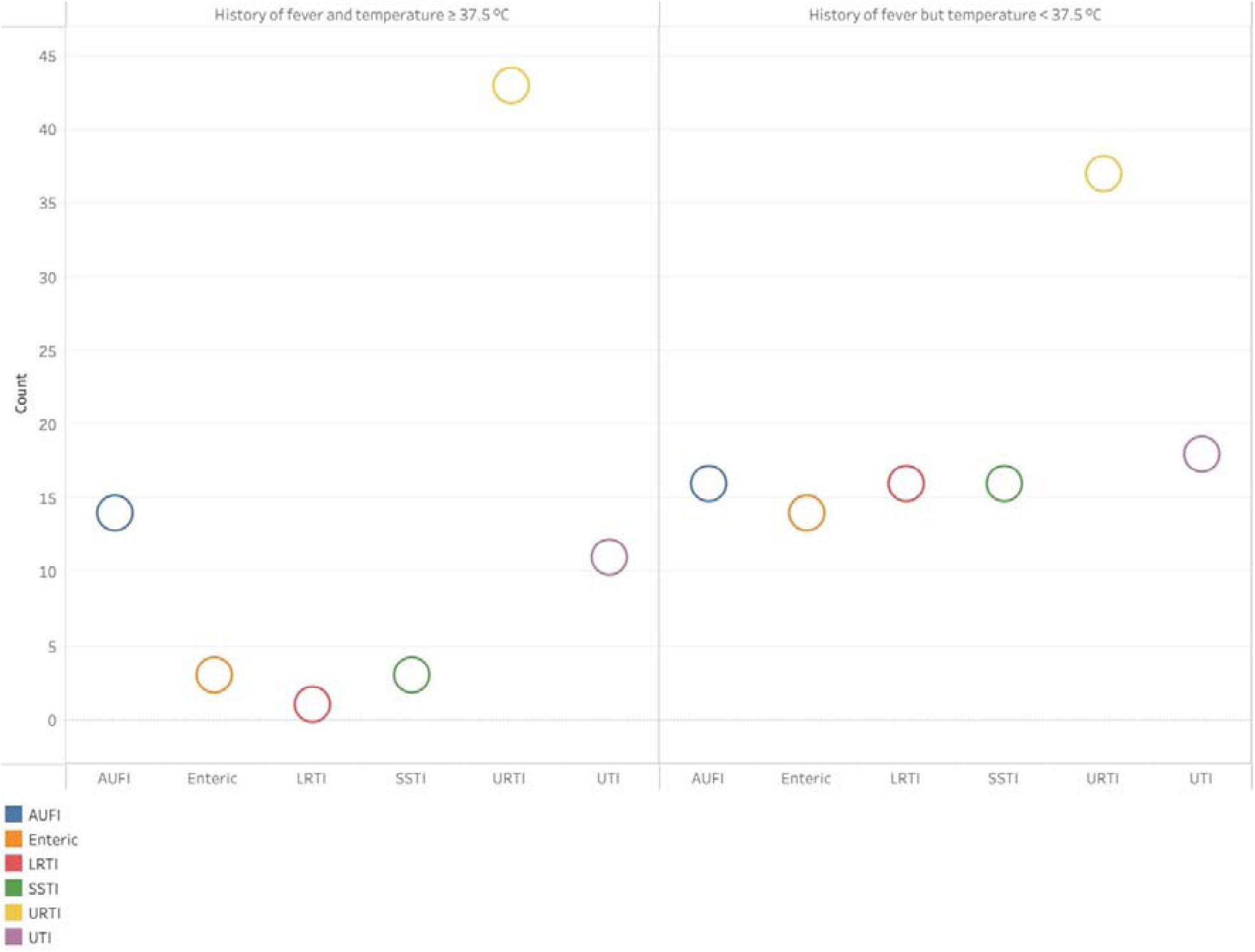
Foci of infection according to the height of temperature mensurated at arrival in febrile patients attending Emergency departments in Rio de Janeiro, Brazil, October 2018-March 2019 AUFI stands for acute undifferentiated febrile illness; LRTI lower respiratory tract infection; SSTI skin and soft tissue infection; URTI upper respiratory tract infection; and UTI urinary tract infection

**Supplementary Figure 4.**
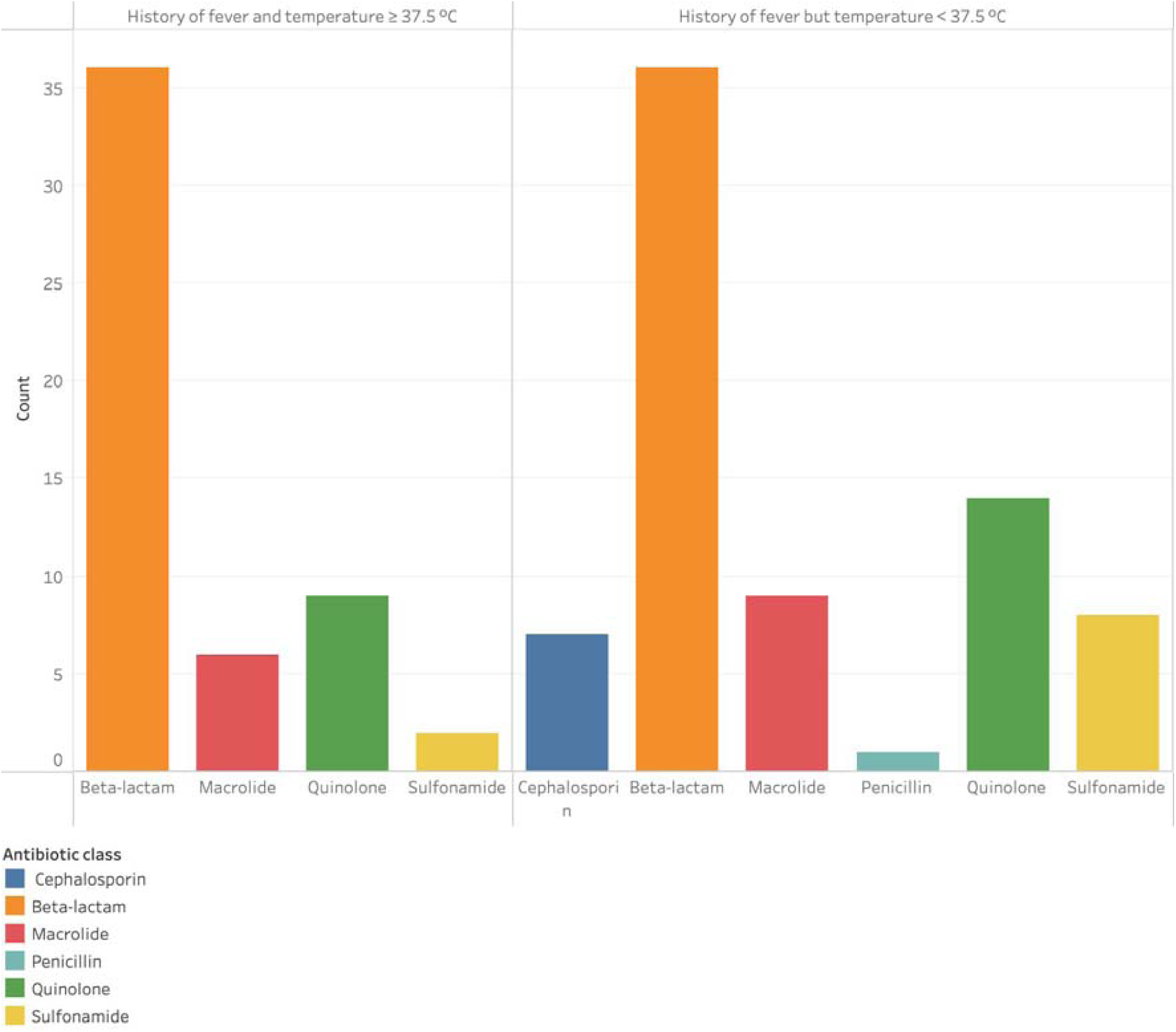
Antibiotic class according to the height of temperature mensurated at arrival in febrile patients attending Emergency departments in Rio de Janeiro, Brazil, October 2018-March 2019

## Key messages

- Fever is a common reason to seek care in patients attending urban Emergency Departments in Rio de Janeiro, Brazil
- Suspicion of infection is the underlying cause of fever in 53% of the febrile subjects, and the respiratory source is the main focus of infection
- The use of antipyretics among febrile patients attending urban Emergency Departments in Rio de Janeiro is ubiquitous, and dipyrone is the antipyretic of choice in our population. However, such use did not influence overt fever presentation in Emergency Departments
- Our findings are essential for designing fever etiology and question the value of adopting a specific temperature cut-off for fever at enrollment, as in many fever studies. We believe that a history of fever as part of the inclusion criteria is a good surrogate of unwellness and might be incorporated systematically in future fever studies.

### Appendix Table of Contents

This supplementary material has been provided by the authors to give readers additional information about their work.

